# Brain-age predicts subsequent dementia in memory clinic patients

**DOI:** 10.1101/2021.04.03.21254781

**Authors:** Francesca Biondo, Amelia Jewell, Megan Pritchard, Dag Aarsland, Claire J. Steves, Christoph Mueller, James H. Cole

## Abstract

**INTRODUCTION:** Research into quantitative neuroimaging biomarkers of dementia risk rarely uses data representative of everyday clinic practice.

**METHODS:** We analysed T1-weighted MRI scans from memory clinic patients (n=1140; 60.2% female and mean [SD] age of 70.0 [10.8] years) to derive ‘brain-age’, an index of age-related brain health. We determined which patients went on to develop dementia (n=476) via linkage to electronic health records.

**RESULTS:** Cox regression indicated a 3% increased risk of dementia per brain-PAD year (brain-PAD = brain-age minus chronological age), *HR*(95% *CI*)=1.03(1.02, 1.04), *p*<0.001, adjusted for age, age^2^, sex, MMSE and normalised brain volume. Brain-PAD remained significant even with a minimum time-to-diagnosis of 3 years (*HR*=1.06) and with MMSE score ≥ 27 (*HR*=1.03).

**DISCUSSION:** Memory clinic patients with older-appearing brains are more likely to receive a subsequent dementia diagnosis. These results from a ‘real-world’ dataset suggest quantitative neuroimaging biomarkers like brain-age could be readily used in the clinic.

**Research in Context:** *SYSTEMATIC REVIEW:* Multiple previous studies were identified that have modelled dementia risk using quantitative neuroimaging, however, screening of participants based on comorbidities and contraindications alongside sociodemographic and healthcare sampling biases, limits the generalisation of these studies to real-world clinical settings. To facilitate better translation from research to the clinic, datasets that are more representative of dementia patient groups are warranted.

*INTERPRETATION:* Brain-age is an index of ‘biological’ age based on a quantitative analysis of T1-weighted MRI scans. Memory clinic patients with biologically older-appearing brains are more likely to receive a subsequent dementia diagnosis, independent of medical history, age, sex, MMSE score and normalised brain volumes. These findings suggest that brain-age has potential to be used early-on in memory clinics as a biomarker to aid detection of patients at high-risk of developing dementia.

*FUTURE DIRECTIONS:* Does the addition of T2-weighted MRI scan information and/or localised brain-age values improve dementia prediction?

## 1 Introduction

The growing global burden of dementia motivates efforts to improve the early identification of people at highest risk. Early dementia-risk identification has important implications for future care planning, the timing of possible interventions and for stratified clinical trial enrolment. Consequently, this growing clinical need potentially warrants a shift of emphasis away from fundamental science underlying dementia to the research into clinical prognosis – the science of predicting the risk of disease [1].

Investigations of predictive models of dementia abound and include cognitive and behavioural markers [2, 3] as well as biomarkers targeting features such as abnormal protein aggregation (e.g., amyloid, tau, neurofilament light) and brain structure, function and metabolism [4, 5]. However, there remains a gap, or “valley of death”, between these basic research findings and their use in the clinic [6, 7]. One common origin of this gap is a deficiency of ecological validity, which refers to both ‘representativeness’ and ‘generalisability’ [8, 9]. A widespread cause of poor ecological validity is selection bias, where participants with certain characteristics are more or less likely to be included in the study than others [10, 11]. The resulting sample misrepresentation limits generalisation to the target population. For example, recruiting patients from a dementia clinic whilst selecting controls from a primary care clinic may lead to potentially erroneous conclusions, such as arthritis and cataracts being more common in controls [12]. As the authors admit, this is likely driven by an asymmetrical representation of non-neurological conditions. Another example is the large prospective cohort study, UK Biobank, which displays a ‘healthy volunteer’ bias whereby the included participants were found to be more health-conscious compared with the general population [13]. Typically, dementia risk prediction studies exclude participants with current or past comorbidities [14-16], which is not representative of most people at risk of dementia [17-19].

Neuroimaging is a strong candidate for predicting dementia, particularly magnetic resonance imaging (MRI) as it is routinely collected in clinical contexts [20]. Previous work has been consistent in reporting brain atrophy [16, 21], thinner cortical thickness [14, 15], microstructure abnormalities including white-matter hyperintensities [22, 23] and differences in functional connectivity [24] as associated to future dementia. One promising approach to predict health outcomes in neurodegenerative diseases is the brain-age paradigm. Brain-age is an index of the brain’s biological age, with previous studies supporting the idea that ‘older’-appearing brains are indicative of a greater risk of age-associated brain diseases and poor health outcomes, including mortality [25-31]. Brain-age has also been associated with subsequent dementia in observational research cohorts [32, 33]. When compared with other AD neuroimaging biomarkers such as CSF-based amyloid and tau markers, or PET markers, brain-age provided an independent contribution in predicting conversion from Mild Cognitive Impairment (MCI) to AD [34].

While promising, these research studies have a key limitation; they are unrepresentative of the general population at-risk for dementia, as research participants are likely to be more highly educated, have a higher IQ, be less ethnically diverse and have fewer comorbidities than the general population [35]. Even in research studies that have gone to great lengths to obtain a representative sample [36], the aforementioned healthy-volunteer bias is challenging to overcome and factors such as the threshold for contraindications for undergoing MRI will differ between research and clinical settings.

In the present study, we sought to improve ecological validity using a large real-world dataset of patients referred to memory clinics for MRI assessment. We hypothesised that brain-age would significantly improve prediction of *if* and *when* memory clinic patients would subsequently develop dementia. This retrospective study analysed structural MRI scans of memory clinic patients whose subsequent presence or absence of dementia was determined via linkage to electronic health records (EHRs). This study was pre-registered (<u>aspredicted.org</u> ref.26262) before data access.

## 2 Method

### 2.1 Participants

This study analysed 1140 memory clinic patients who were referred for a neuroimaging assessment at the South London and the Maudsley (SLaM) NHS trust. SLaM is one of the largest secondary mental healthcare providers in Europe, serving over 1.36 million residents from predominantly four London boroughs (Croydon, Lambeth, Lewisham and Southwark) [37, 38]. BRCMEM and BRCDEM are ongoing studies and the oldest scan date in our sample is 28/01/2011.

Patients’ demographic and clinical data were available from de-identified electronic health records (EHRs), which were accessed via the Clinical Record Interactive Search (CRIS) based at the BRC [37-39]. CRIS is a clinical database with a robust data governance framework with ethical approval for secondary data analysis (Oxford REC C reference 18/SC/0372). CRIS provided linkage between EHRs and the BRCMEM and BRCDEM neuroimaging data, as well as linkage to two other datasets, the Hospital Episode Statistics (HES) and the Office of National Statistics (ONS). HES is a national dataset that contains data of outpatient appointments and hospital admissions at NHS hospitals in England [40]. ONS contains the date and cause of death for all deaths registered in England and Wales [41]. HES and ONS were used to supplement the EHRs with diagnostic and mortality data.

Permission to access EHRs, HES, ONS, BRCMEM and BRCDEM databases was granted via CRIS (reference 19-008) and accessed on 11/10/2019. Informed consent for research-use of neuroimaging data was originally obtained from the BRCMEM and BRCDEM participants at time of assessment. Data preparation was carried out via two routes, clinical and neuroimaging (**Figure 1)**. The analyses code will be made available here: https://github.com/biondof/BARCODE.

**Figure 1.**
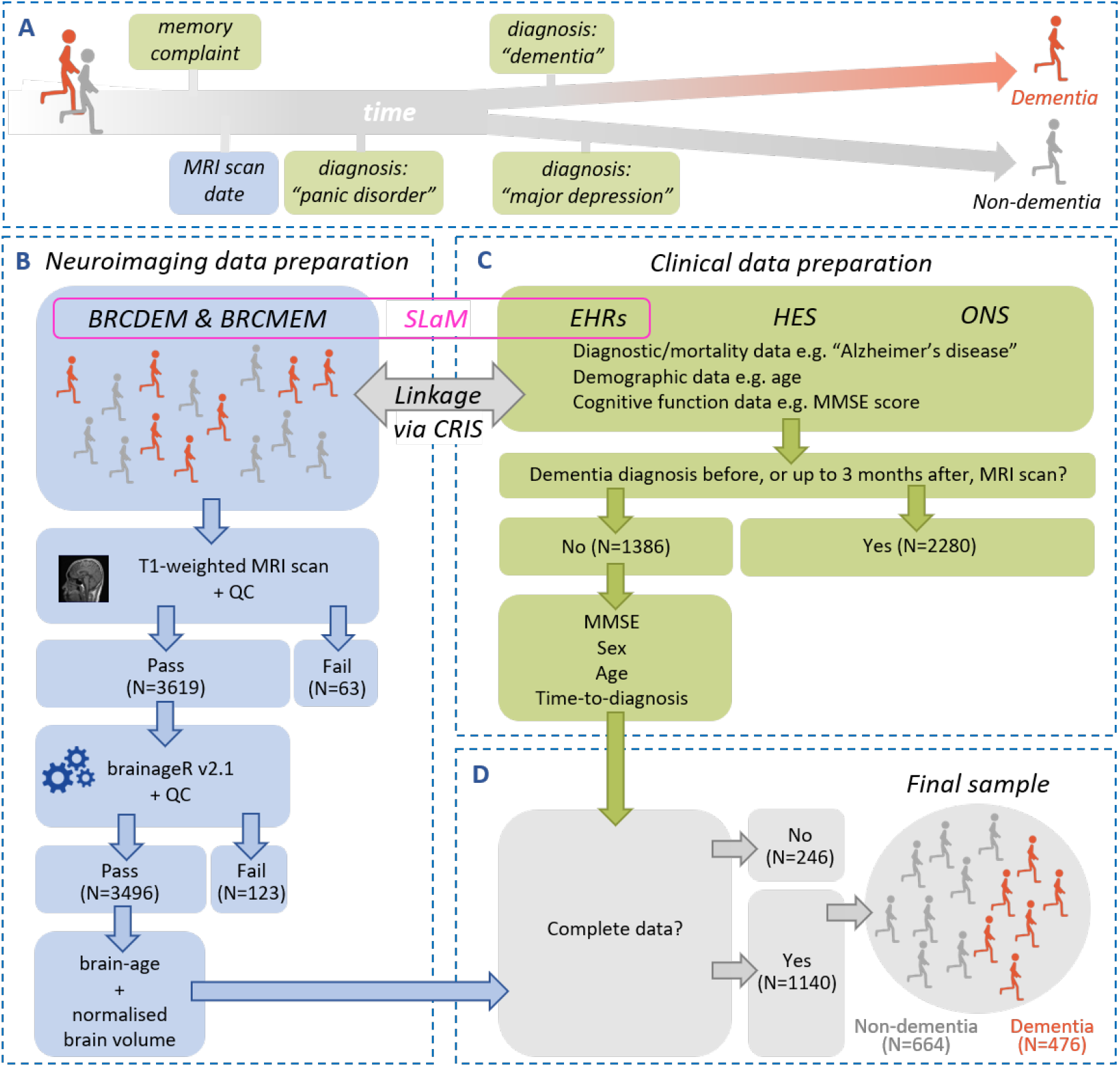
Data pre-processing pipeline. **(A)** Memory clinic patients from the BRCDEM and BRCMEM studies were classified into two types: dementia (orange) or non-dementia (grey) **(B) Neuroimaging data preparation**: BrainageR v2.1 software was applied to T1-weighted MRI scans to obtain a brain-age estimate and normalised brain volume for each patient. Quality Control (QC) aimed to remove cases with image artefacts and occurred at two stages, before and after segmentation. **(C) Clinical data preparation:** clinical data was retrieved across three databases, EHRs, HES and ONS which were accessed and linked to the neuroimaging data via CRIS (Clinical Record Interactive Search). These databases provided diagnostic, demographic, cognitive and mortality data. This allowed us to determine which patients went on to get, or not, a dementia diagnosis in their medical journey allowing labelling into dementia or non-dementia types. Patients who were diagnosed with dementia before, or up to 3 months after the neuroimaging assessment, were excluded from our analyses. **(D) Merging the neuroimaging and clinical data**: the final dataset (N=1140) included only complete cases for the following variables: sex, age, brain-age, normalised brain volume, MMSE (Mini Mental State Examination) and scanner information. BRCDEM, BRCMEM: names of two clinical-research studies for memory clinic patients based and part of routine clinical care at SLaM (South London and Maudsley Hospital); EHRs (Electronic Health Records); HES (Hospital Episodes Statistics); ONS (Office of National Statistics); CRIS (Clinical Record Interactive Search).

### 2.2 Clinical data preparation

The CRIS-enabled linkage of neuroimaging data to EHRs, identified a total of 3666 patients. Of these, 2472 were classified as dementia patients and 1194 as non-dementia patients. Classification was achieved by extraction of diagnostic information and/or cause of death across EHRs. A dementia diagnosis was operationalised as a positive search result for the following terms: ICD-10 codes F00-F03, G30-G32, “dementia”, “Alzheimer’s disease”, “Alzheimer” and “Lewy”. The search was carried out in both structured and unstructured fields of the EHRs. Unstructured fields included free-text clinical notes and the search was achieved via natural language processing (NLP) applications developed within CRIS at the BRC [38, 39]. A negative search result classified the patient as a non-dementia type.

For the dementia patients, the diagnosis time was the date of the first instance of a dementia diagnosis after the neuroimaging assessment. For the non-dementia patients, the diagnosis time was the last instance of a diagnostic clinical entry after the neuroimaging assessment. A dementia diagnosis was detected before the neuroimaging assessment in 745 dementia patients (suggesting the diagnosis had been made based on clinical assessment after referral to the MRI unit, but before the actual scan); these were excluded. Seventy-two non-dementia cases were excluded because their most recent non-dementia diagnosis preceded the neuroimaging assessment.

We did not exclude anyone based on their medical histories (except dementia, as previously described). This deviates from our pre-registered statement because, in retrospect, we opted to better capture the heterogeneity of memory clinic patients as this would increase ecological validity.

As the time between diagnosis and neuroimaging assessment narrows, the predictive value of the MRI scan becomes more redundant. To address this, we set a minimum threshold of 3 months between neuroimaging assessment and diagnosis, based on SLaM NHS clinical guidelines of the maximum wait for a diagnosis. Thresholding reduced the sample from 2848 to 1386.

In addition to diagnostic information, the following variables ascertained at, or closest to, the neuroimaging assessment were obtained from the EHRs: age, sex, Mini-Mental State Examination (MMSE) and Addenbrookes’ Cognitive Examination (ACE). The MMSE and ACE are brief cognitive functioning tests commonly used in the clinic and helpful at detecting dementia [42, 43]. The scores range from 0-30 and 0-100, respectively, with low scores indicating poor cognition. NLPs were used to extract MMSE and ACE scores from both structured and unstructured EHRs fields.

### 2.3 Neuroimaging data preparation

In total, 3682 T1-weighted MRI scans were accessed via the BRCDEM and BRCMEM studies. These used similar but not identical acquisition parameters (see **Supplementary Materials**). Visual quality control (QC) was conducted to detect image artefacts; 63 scans were excluded due to poor quality such as motion artefacts and field inhomogeneities.

Brain-age was calculated using brainageR (v2.0), an open-access software designed to generate brain-predicted age from raw T1-weighted MRI scans (https://github.com/james-cole/brainageR)[29]. The brainageR software involves two main stages, pre-processing and prediction. In the pre-processing stage, the images were segmented and normalised via SPM12 software (www.fil.ion.ucl.ac.uk/spm/software/spm12/). PNGs of these processes were generated via the slicesdir FSL function that facilitated a second visual QC to verify segmentation accuracy; 123 images were excluded due to gross segmentation errors. Normalised images were loaded into R *[44]* and vectorised. GM, WM and CSF vectors were masked using a 0.3-thresholded average image template based on the brainageR’s model training dataset and then combined.

In the prediction stage, the brainageR model was applied to the vectorised and masked study images to estimate a brain-age score for each. BrainageR had been previously trained to predict age from normalised brain volumetric maps of n=3377 healthy individuals from seven publicly-available datasets using a Gaussian Processes Regression. Using principal component analysis, the top principal components capturing 80% of the variance in age were retained. The resulting rotation matrix for 435 principal components was then applied to the new imaging data to predict age. Model performance (Pearson’s correlation between chronological age and brain-predicted age, *r*; mean absolute error, MAE) for internal and external validation was: n=857, *r* = 0.973, MAE = 3.933 years and n=611, *r* = 0.947, MAE = 4.90 years, respectively.

For each image, the final output of brainageR was a brain-predicted age value with 95% confidence intervals. Brain-predicted age difference (brain-PAD) was calculated by subtracting chronological age from brain-predicted age. Volumetric measures of GM, WM and CSF were generated by SPM Segment. Normalised brain volume was calculated as the sum of GM and WM volumes, divided by the sum of GM, WM and CSF volumes.

Neuroimaging data were subsequently merged with the clinical data. Out of 1386 cases, 81 were removed because of a missing brain-age score or a different MRI scanner and 165 were removed because of missing MMSE scores. The final sample (n=1140) consisted of 664 dementia and 476 non-dementia patients.

### 2.5 Statistical analyses

To examine whether brain-age was associated with subsequent dementia diagnosis (at least 3 months later), we ran two analyses. First, we used logistic regression to test whether baseline brain-PAD was associated with future dementia status (dementia versus non-dementia). Secondly, we conducted a survival analysis using Cox proportional hazards regression to determine whether baseline brain-PAD predicted time-to-dementia. Cox regression allows for the predictive modelling of the time-to-event (time-to-dementia), also referred to as survival time or event time. For the non-dementia patients, time-to-event was right-censored using the data of the most recent diagnostic clinical entry. For both analyses, the predictor of interest was brain-PAD and covariates were sex, age, age^2^, MMSE score and normalised brain volume. Age and age^2^ were included to address the bias due to the correlation between chronological age and brain-PAD [45]. The problem of multicollinearity between age and age^2^ was minimized by using their polynomial terms.

Next, we ran two sensitivity analyses by subsetting the sample data and re-running both logistic and cox regression analyses. Firstly, we examined whether brain-PAD was predictive of dementia over a longer period, using a minimum duration of 3 years between neuroimaging assessment and diagnosis. Secondly, we tested whether brain-PAD was predictive of dementia in patients appearing to be largely cognitively unimpaired, by excluding those with MMSE scores below 27 [46].

## 3 Results

Baseline characteristics of the final sample (n=1140) are described in **Table 1**. At the time of neuroimaging assessment, 60.18% were female, the mean age was 69.99 (SD 10.80) years, the mean MMSE score was 23.01 (SD 6.90), the mean ACE score was 72.30 (SD 15.63) and the mean normalised brain volume was 0.7239 (SD 0.0591) litres. Compared with the non-dementia group, the dementia group had more females, was older, had lower MMSE and ACE scores and had smaller brain volumes.

**Table 1.** Sample Characteristics.

| Group | All | Non-dementia | Dementia | Non-dem<br>vs. Dem<br>(*p*-value) |
| --- | --- | --- | --- | --- |
| **N** | 1140 | 664 | 476 |  |
| **Female %** | 60.18 | 57.68 | 63.66 | 0.0422 |
| **Age** |  |  |  |  |
| Complete cases % | 100.00 | 100.00 | 100.00 |  |
| Mean (SD) | 69.99 (10.80) | 66.95 (11.12) | 74.24 (8.74) | <0.0001 |
| Median | 71.00 | 68.00 | 75.00 |  |
| Min-Max | 27.00-95.00 | 27.00-95.00 | 42.00-95.00 |  |
| **MMSE** |  |  |  |  |
| Complete cases % | 100.00 | 100.00 | 100.00 |  |
| Mean (SD) | 23.01 (6.90) | 23.88 (6.59) | 21.79 (7.15) | <0.0001 |
| Median | 26.00 | 26.00 | 24.00 |  |
| Min-Max | 0.00-30.00 | 1.00-30.00 | 0.00-30.00 |  |
| **Time-to-Event** |  |  |  |  |
| Complete cases % | 100.00 | 100.00 | 100.00 |  |
| Mean (SD) | 1.86 (1.61) | 2.18 (1.66) | 1.41 (1.42) |  |
| Median | 1.29 | 1.70 | 0.81 |  |
| Min-Max | 0.25-7.81 | 0.25-7.81 | 0.25-7.49 |  |
| **ACE** |  |  |  |  |
| Complete cases % | 35.09 | 39.31 | 29.20 |  |
| Mean (SD) | 72.30 (15.63) | 74.81 (14.82) | 67.58 (16.05) | <0.0001 |
| Median | 75.00 | 78.00 | 70.00 |  |
| Min-Max | 22.00-99.00 | 26.00-99.00 | 22.00-96.00 |  |
| **Normalised brain volume** |  |  |  |  |
| Complete cases % | 100.00 | 100.00 | 100.00 |  |
| Mean (SD) | 0.7239 (0.0591) | 0.7405 (0.0571) | 0.7009 (0.0538) | <0.0001 |
| Median | 0.7261 | 0.7453 | 0.7073 |  |
| Min-Max | 0.4728-0.8800 | 0.4738-0.8800 | 0.4728-0.8633 |  |

*SD = Standard Deviation; MMSE = Mini-Mental State Examination score; normalised brain volume is measured in litres.*

The median time-to-dementia diagnosis was 0.81 years (interquartile range 0.41-1.92). Associations between brain-PAD and outcomes of dementia diagnosis (logistic regression) and time-to-dementia diagnosis (Cox regression) are presented in **Table 2**. A higher brain-PAD was significantly associated with a dementia diagnosis in both the logistic regression: odds ratio (OR) = 1.03 (CI 1.02-1.05), and the Cox regression: hazards ratio (HR) = 1.03 (CI 1.02-1.04). Age, sex, MMSE and normalised brain volume, but not age^2^, were also significantly associated with outcome (see **Supplementary Materials**). These results indicated that while keeping the covariates constant, every +1 year of brain-PAD was accompanied with a 3% relative increased risk of a dementia diagnosis (*p*<0.0001), both when considering time-to-dementia (Cox regression) or dementia/non-dementia status (logistic regression) (**Figure 2**).

**Table 2.** Association of brainPAD (adjusted for covariates) with incident dementia assessed by logistic regression and Cox proportional hazards models, in the total study sample and in subsamples based on sensitivity analyses. *The table above shows results from three analyses: the main analysis using the total study sample (N=1140); sensitivity analysis I (N=249) which included patients that were diagnosed at least 3 years after the neuroimaging assessment and, sensitivity analysis II (N=471) which included only a subsample of patients who scored 27 or more on the MMSE at the time of the neuroimaging assessment. OR = odds ratio; HR = hazards ratio*

|  | n/N | Logistic regression |  |  |  | Cox regression |  |  |  |
| --- | --- | --- | --- | --- | --- | --- | --- | --- | --- |
|  |  | OR | (95% CI) | p-value |  | HR | (95% CI) | p-value |  |
| **Main analysis** | 476/1140 | 1.03 | (1.02-1.05) | <0.0001 | *** | 1.03 | (1.02-1.04) | <0.0001 | *** |
| **Sensitivity analysis I** | 66/249 | 1.06 | (1.03-1.10) | 0.0011 | ** | 1.06 | (1.02-1.09) | <0.0001 | *** |
| **Sensitivity analysis II** | 146/471 | 1.02 | (1.00-1.05) | 0.0544 |  | 1.03 | (1.01-1.05) | <0.0001 | *** |

OR= Odds Ratio; CI= Confidence Intervals; number of dementia cases (n); total number of participants (N)
\*\*\* $p$ -value<0.001, \*\* $p$ -value<0.01
Sensitivity analysis I= minimum 3 years for time-to-diagnosis; Sensitivity analysis II= MMSE score $\geq 27$

**Figure 2.**
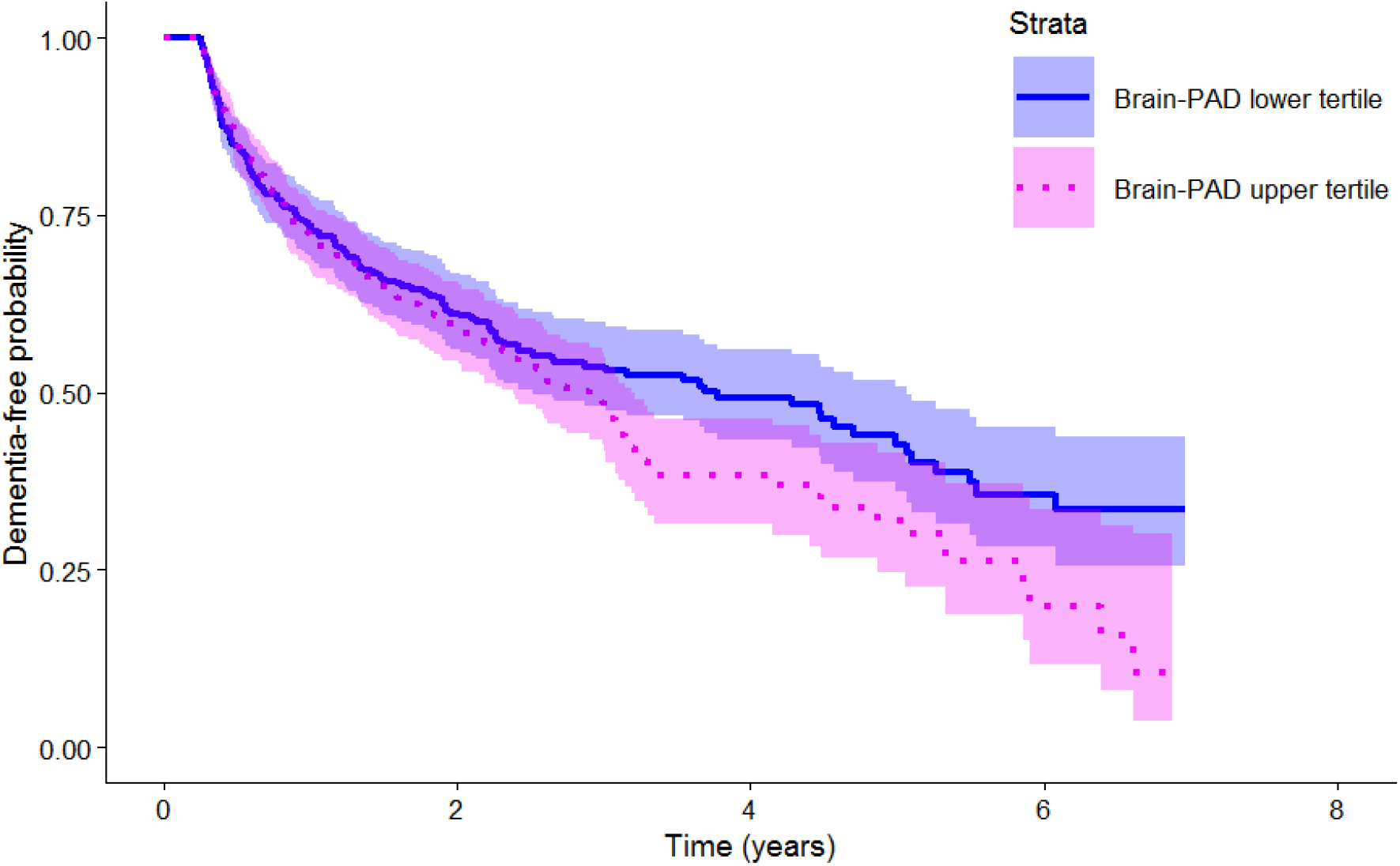
depicts a Kaplan-Meier plot illustrating the probability of being dementia-free based on brainPAD score. At the time of the neuroimaging assessment (time 0 on the x-axis), all patients are dementia-free. Over time, patients with higher brainPAD scores (pink) are more likely to get a dementia diagnosis and more rapidly than the ones with lower brainPAD scores (blue).

Assumptions for both regression models were met. For the logistic regression, the residuals were normally distributed and for the Cox regression, the standardised residuals of the covariates were not correlated to event time indicating proportional hazards (χ^2^(6)=11.5, *p*=0.074).

Congruent with our main results, the non-dementia group’s brain-age showed a relatively smaller deviation from their chronological age compared with the dementia group, as illustrated in **Figure 3**, with the line of best fit closely aligned to the identity line in green (left panel) but less so for the dementia group (right panel). Also, the deviation negatively correlated with age, likely driven by a regression-to-the-mean effect, hence we accounted for this in our analyses by covarying age and age^2^ [45].

**Figure 3.**
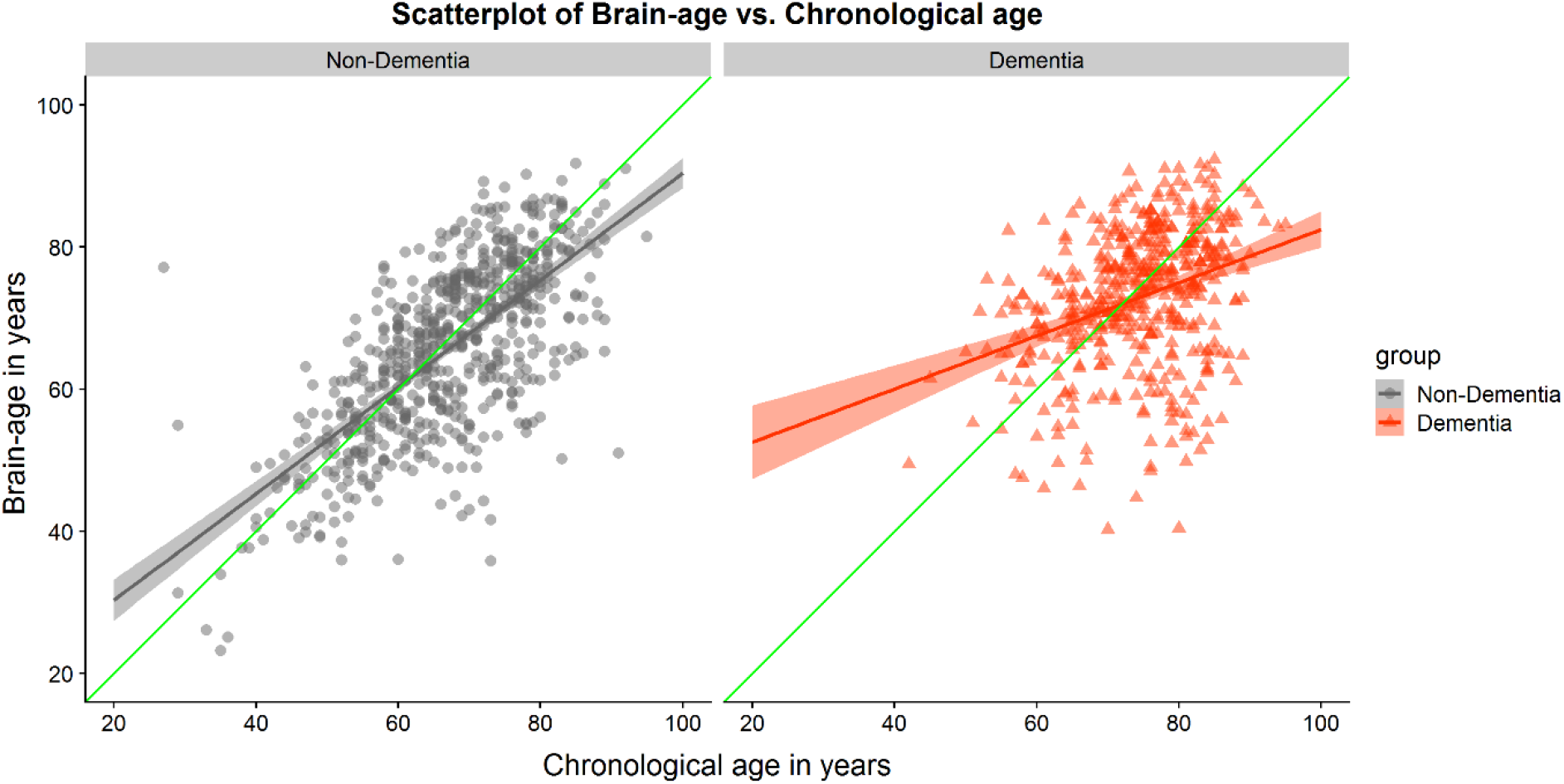
The scatterplots show chronological age (x-axis) and brain-age (y-axis) split by group, non-dementia (grey) in the left panel and dementia (orange) in the right panel. The identity line (green) shows the ideal case when chronological age matches the brain-age estimate, x=y. Lines of best fit (orange, grey) within each group are both positive showing that brain-age estimates tend to be larger than chronological age for both groups of patients and in particular for the dementia group.

The first sensitivity analysis restricted to patients with a time-to-diagnosis minimum of 3 years (n=249) resulted in an OR of 1.06 (CI 1.03-1.10) and HR of 1.06 (CI 1.02-1.09), which were both statistically significant. This indicated that for every +1 year of brain-PAD there was a 6% relative increased risk of dementia, even when the neuroimaging assessment preceded diagnosis by at least 3 years. The second sensitivity analysis restricted to patients with baseline MMSE scores of above 26 (n=471), resulted in an HR of 1.03 (CI 1.01-1.05). This indicated a 3% increased risk of dementia for every +1 brain-PAD year, even when cognition was not clearly impaired. The latter analysis was statistically significant for the Cox regression but not using logistic regression. For other covariates’ OR and HR, see **Supplementary Materials**.

## 4 Discussion

In this study, we hypothesised that brain-age, a structural MRI-based biomarker, would predict subsequent dementia in a real-world sample of memory clinic patients. We found that every additional year of brain-PAD incurred a 3% relative increased risk of dementia. This implies that memory patient clinics with older-looking brains are more likely to receive a subsequent dementia diagnosis, independent of sex, baseline age, MMSE score and normalised brain volume. Accordingly, a patient with +5 brain-PAD years has 15% added risk of dementia compared to one with 0 brain-PAD years, whilst keeping the covariates equal (e.g., both are female 65-year olds with similar MMSE scores and normalised brain volumes). Moreover, sensitivity analysis I revealed that this risk increased to 6% when the interval between neuroimaging assessment and dementia diagnosis was increased from a minimum of 3 months to at least 3 years. This could indicate that neuroimaging biomarkers are more informative in people further away from the development of manifest clinical symptoms of dementia.

Our findings are consistent with previous studies that used the brain-age paradigm to predict subsequent dementia [32, 33]. For example, the Wang *et al*. [33] study showed that every year of brain-PAD had a 9% increased risk of incident dementia, even when assessment preceded diagnosis by 5 years. Also, the Gaser *et al*. [32] study reported a 10% increased risk in converting from MCI to AD with a 3-year follow-up. Some important differences exist between our study and these previous reports, particularly regarding sample characteristics. In Wang and colleagues’ study, the sample was a population-based cohort in which participants who developed dementia were compared to those who did not and hence the latter are likely to have included many healthy participants. Instead, all our controls were patients who were referred to a memory clinic and who did eventually get a diagnosis, often a psychiatric or neurological one. Hence, the case-control contrast in the Wang’s study is expected to be larger than ours given that patients with psychiatric and neurological disorders are reported to show larger brain-PADs than healthy participants [25, 26, 30]. In our study, brain-PAD remained predictive despite our sample including co-morbidities and complex medical histories.

Another important finding came from our sensitivity analysis II; when cognitive functioning was unimpaired or at least not clearly impaired (MMSE ≥ 27), brain-PAD remained a significant predictor of the disease. It is precisely when routine assessments in the memory clinic yield ambiguous results that the clinician may consider other more invasive or expensive tests to aid decision-making. This further illustrates the potential role of quantitative neuroimaging biomarkers in the clinic for augmenting clinical decision-making. For example, it is not uncommon for memory clinic patients with poor cognition to receive ‘age appropriate’ radiological report on an MRI scan. Quantitative neuroimaging analysis could offer the possibility to reconcile such findings.

One major strength of this study is having used data from pre-existing standard memory clinic workflows rather than by experimental, and hence, artificial design. This confers two main advantages. The first is added ecological validity. This means that our findings are more representative of our target population - memory clinic patients. Our sample is drawn from south-east London and characterised by a highly diverse ethnic background, which is an asset given the widespread criticism for its scarcity. Notably, south-east London is not fully representative of nationwide memory clinic patients [38] but still offers an improvement over some sources of misrepresentation such as selection bias.

Secondly, it illustrates the feasibility of implementing clinical predictive models involving quantitative neuroimaging biomarkers, which could be automatically integrated with other clinical data (e.g., radiology reports or EHRs). Connecting neuroimaging biomarkers like brain-age with EHRs has broad potential; it could aid the memory clinician with prognosis and support clinical trials including stratification, staging of disease severity and outcome markers [5] and, neuropsychiatric research at large [26, 28, 30].

To put our results into context we considered how the effect size of the risk conferred by brain-PAD compared to common dementia risk factors. The study by Gottesman *et al*.[47] reports HR(CI) for incident dementia of 1.14 (0.99-1.31), 1.39 (1.22-1.59) and 1.77 (1.53-2.04) respectively for obesity, hypertension and diabetes relative to normal healthy conditions. Comparing these to brain-PAD is not straightforward considering study differences such as sample characteristics and covariate adjustments. However, if we hypothetically put set aside these caveats, and converted these HRs to brain-PAD years based on our model, this would give +4.7, +11.8 and +20.5 brain-PAD years, respectively. Broadly speaking, this would mean that someone with a brain looking 11.8 years older than their chronological age would have a similar long-term risk of incident dementia to someone classified as having hypertension and 0 brain-PAD years.

Various limitations of this study should be acknowledged. The first pertains to false negatives and false positives. The non-dementia group was defined on a negative search result for a dementia diagnosis, however, it is possible that some developed dementia that was unrecorded. Nevertheless, if our sample did contain such false negatives then, the contrast between dementia versus non-dementia would reduce and consequently, our current finding would reflect a more conservative version than its true value. False positives - mistaken cases of dementia patients - are less likely though not impossible, despite constant efforts at improving validation of information extracted from EHRs [38, 39]. Future efforts in this direction could include enhanced triangulation of observations by linking EHRs to additional databases, such as general practitioner records. A second limitation pertains to the precision of the timestamps of EHRs events given the lack of consistency concerning whether the date reflects the time of the clinical event or the time the clinical event was recorded on the system. However, we assume that this inconsistency would exist at random across groups and not by a large interval. Thirdly, brain-age was calculated using only one modality of structural MRI scans. Although a T1-weighted brain-age model has been extensively validated across many studies it leaves a rich resource of common clinical scans, T2-weighted ones, unused. A multimodal brain-age model could improve the utility of this biomarker [48]. Finally, although part of the appeal of the brain-age model lies in the simplicity of multivariate-to-univariate transformation (i.e., many voxels reduced to a single brain-age value), ongoing developments in brain-age modelling provide localised brain-age estimates [49] that could allow for novel composite brain-age scores of brain regions known to be critical to dementia (e.g., temporal lobe versus whole brain).

Overall, our findings demonstrate the value of using brain-age as a sensitive biomarker that has the potential to be used early-on in memory clinics to detect patients at high risk of developing dementia. This ‘real-world’ study further paves the way for the role of quantitative neuroimaging in bridging the gap between basic research and clinical applications, in particular, prognostication and stratification of dementia syndromes.

## Data Availability

Patient data not available.

## Acknowledgements and Conflicts of Interest

The study was funded by a Wellcome Trust Seed Award in Science (213996/Z/18/Z). CM receives salary support from the National Institute for Health Research (NIHR) Biomedical Research Centre at South London and Maudsley NHS Foundation Trust and King’s College London. CJS receives salary support from Medical Research Council, the Wellcome Trust and the Chronic Disease Research Foundation. This paper represents independent research partly funded by the National Institute for Health Research (NIHR) Biomedical Research Centre at South London and Maudsley NHS Foundation Trust and King’s College London. The views expressed are those of the authors and not necessarily those of the NHS, the NIHR or the Department of Health and Social Care. Authors report no conflicts of interest. *Potential conflict of interest:* JHC is a scientific advisor to and shareholder in Brain Key, a medical image analysis company.

## Acronyms

(ACE): Addenbrookes’ Cognitive Examination
(AD): Alzheimer’s disease
(brain-PAD): Brain Predicted Age Difference
(BRC): Maudsley NIHR Biomedical Research Centre
(CRIS): Clinical Record Interactive Search
(EHRs): Electronic Health Records
(HES): Hospital Episode Statistics
(MCI): Mild Cognitive Impairment
(MMSE): Mini-Mental State Examination
(MRI): Magnetic Resonance Imaging
(NLP): Natural Language Processing
(ONS): Office of National Statistics
(PET): Positron Emission Tomography
(QC): Quality Control
(SLaM NHS): South London and the Maudsley National Health Service Foundation Trust

## Notes

### Author Declarations

Oxford REC C reference 18/SC/0372 & NIHR Maudsley Biomedical Research Centre: CRIS reference 19-008

## References

1. Fusar-Poli, P., et al., The Science of Prognosis in Psychiatry: A Review. JAMA Psychiatry, 2018. 75(12): p. 1289-1297.

2. Belleville, S., et al., Neuropsychological Measures that Predict Progression from Mild Cognitive Impairment to Alzheimer’s type dementia in Older Adults: a Systematic Review and Meta-Analysis. Neuropsychology Review, 2017. 27(4): p. 328–353.

3. Montero-Odasso, M.M., et al., Association of Dual-Task Gait With Incident Dementia in Mild Cognitive Impairment: Results From the Gait and Brain Study. JAMA Neurology, 2017. 74(7): p. 857–865.

4. Humpel, C., Identifying and validating biomarkers for Alzheimer’s disease. Trends in Biotechnology, 2011. 29(1): p. 26–32.

5. Jack, C.R., et al., NIA-AA Research Framework: Toward a biological definition of Alzheimer’s disease. Alzheimer’s and Dementia, 2018. 14(4): p. 535–562.

6. Gamo, N.J., et al., Valley of death: A proposal to build a “translational bridge” for the next generation. Neuroscience Research, 2017. 115: p. 1–4.

7. Cole, J.H., Neuroimaging Studies Illustrate the Commonalities Between Ageing and Brain Diseases. BioEssays, 2018. 40(7).

8. Burgess, P.W., et al., The case for the development and use of “ecologically valid” measures of executive function in experimental and clinical neuropsychology. Journal of the International Neuropsychological Society, 2006. 12(2): p. 194–209.

9. Kvavilashvili, L. and J. Ellis, Ecological validity and the real-life/laboratory controversy in memory research: A critical (and historical) review. History & Philosophy of Psychology, 2004. 6: p. 59–80.

10. Simundić, A.-M., Bias in research. Biochemia medica, 2013. 23(1): p. 12–15.

11. Prince, M., 9 - Epidemiology, in Core Psychiatry (Third Edition), P. Wright, J. Stern, and M. Phelan, Editors. 2012, W.B. Saunders: Oxford. p. 115–129.

12. Broe, G.A., et al., A case-control study of alzheimer’s disease in australia. Neurology, 1990. 40(11): p. 1698–1707.

13. Fry, A., et al., Comparison of Sociodemographic and Health-Related Characteristics of UK Biobank Participants with Those of the General Population. American Journal of Epidemiology, 2017. 186(9): p. 1026–1034.

14. Dickerson, B.C., et al., Alzheimer-signature MRI biomarker predicts AD dementia in cognitively normal adults. Neurology, 2011. 76(16): p. 1395–1402.

15. Li, Y., et al., Discriminant analysis of longitudinal cortical thickness changes in Alzheimer’s disease using dynamic and network features. Neurobiology of Aging, 2012. 33(2): p. 427.e15-427.e30.

16. Vemuri, P., et al., Serial MRI and CSF biomarkers in normal aging, MCI, and AD. Neurology, 2010. 75(2): p. 143–151.

17. Bunn, F., et al., Comorbidity and dementia: A scoping review of the literature. BMC Medicine, 2014. 12(1).

18. Livingston, G., et al., Dementia prevention, intervention, and care: 2020 report of the Lancet Commission. The Lancet, 2020. 396(10248): p. 413–446.

19. Browne, J., et al., Association of comorbidity and health service usage among patients with dementia in the UK: A population-based study. BMJ Open, 2017. 7(3).

20. Rathore, S., et al., A review on neuroimaging-based classification studies and associated feature extraction methods for Alzheimer’s disease and its prodromal stages. NeuroImage, 2017. 155: p. 530–548.

21. Plant, C., et al., Automated detection of brain atrophy patterns based on MRI for the prediction of Alzheimer’s disease. NeuroImage, 2010. 50(1): p. 162–174.

22. Prins, N.D. and P. Scheltens, White matter hyperintensities, cognitive impairment and dementia: An update. Nature Reviews Neurology, 2015. 11(3): p. 157–165.

23. Zeestraten, E.A., et al., Change in multimodal MRI markers predicts dementia risk in cerebral small vessel disease. Neurology, 2017. 89(18): p. 1869–1876.

24. Hohenfeld, C., C.J. Werner, and K. Reetz, Resting-state connectivity in neurodegenerative disorders: Is there potential for an imaging biomarker? NeuroImage: Clinical, 2018. 18: p. 849–870.

25. Cole, J.H. and K. Franke, Predicting Age Using Neuroimaging: Innovative Brain Ageing Biomarkers. Trends in Neurosciences, 2017. 40(12): p. 681–690.

26. Kaufmann, T., et al., Common brain disorders are associated with heritable patterns of apparent aging of the brain. Nature Neuroscience, 2019. 22(10): p. 1617–1623.

27. Han, L.K.M., et al., Brain aging in major depressive disorder: results from the ENIGMA major depressive disorder working group. Molecular Psychiatry, 2020.

28. Franke, K. and C. Gaser, Ten Years of BrainAGE as a Neuroimaging Biomarker of Brain Aging: What Insights Have We Gained? Frontiers in Neurology, 2019. 10.

29. Cole, J.H., et al., Brain age predicts mortality. Molecular Psychiatry, 2018. 23(5): p. 1385–1392.

30. Cole, J.H., et al., Brain age and other bodily ‘ages’: implications for neuropsychiatry. Molecular Psychiatry, 2019. 24(2): p. 266–281.

31. Cole, J.H., et al., Longitudinal Assessment of Multiple Sclerosis with the Brain-Age Paradigm. Annals of Neurology, 2020. 88(1): p. 93–105.

32. Gaser, C., et al., BrainAGE in Mild Cognitive Impaired Patients: Predicting the Conversion to Alzheimer’s Disease. PLoS ONE, 2013. 8(6).

33. Wang, J., et al., Gray Matter Age Prediction as a Biomarker for Risk of Dementia. Proceedings of the National Academy of Sciences, 2019. 116(42): p. 21213.

34. Popescu, S.G., et al., Nonlinear biomarker interactions in conversion from mild cognitive impairment to Alzheimer’s disease. Human Brain Mapping, 2020. n/a(n/a).

35. Petersen, R.C., et al., Alzheimer’s Disease Neuroimaging Initiative (ADNI): Clinical characterization. Neurology, 2010. 74(3): p. 201–209.

36. Ikram, M.A., et al., The Rotterdam Scan Study: design and update up to 2012. European Journal of Epidemiology, 2011. 26(10): p. 811–824.

37. Stewart, R., et al., The South London and Maudsley NHS Foundation Trust Biomedical Research Centre (SLAM BRC) case register: development and descriptive data. BMC Psychiatry, 2009. 9(1): p. 51.

38. Perera, G., et al., Cohort profile of the South London and Maudsley NHS Foundation Trust Biomedical Research Centre (SLaM BRC) Case Register: Current status and recent enhancement of an Electronic Mental Health Record-derived data resource. BMJ Open, 2016. 6(3).

39. NIHR BRC. NIHR Maudsley BRC: Natural Language Processing (NLP) Service. 2021 [cited 2021 12/02/2021]; Available from: https://www.maudsleybrc.nihr.ac.uk/facilities/clinical-record-interactive-search-cris/cris-natural-language-processing/.

40. Academy of Medical Royal Colleges, Hospital_Episode_Statistics_quality_value_data_0511. 2011.

41. NHS Digital, Primary care mortality database. 2019.

42. Burns, A., C. Brayne, and M. Folstein, Mini-Mental State: A practical method for grading the cognitive state of patients for the clinician. M. Folstein, S. Folstein and P. McHugh, Journal of Psychiatric Research (1975) 12, 189-198. International Journal of Geriatric Psychiatry, 1998. 13(5): p. 285–294.

43. Hodges, J.R. and A.J. Larner, Addenbrooke’s cognitive examinations: ACE, ACE-R, ACE-III, ACEapp, and M-ACE, in Cognitive Screening Instruments: A Practical Approach. 2016. p. 109–137.

44. R Core Team, R: A Language and Environment for Statistical Computing, kin R Foundation for Statistical Computing. 2019: Vienna, Austria.

45. de Lange, A.-M.G. and J.H. Cole, Commentary: Correction procedures in brain-age prediction. NeuroImage: Clinical, 2020. 26: p. 102229.

46. O’Bryant, S.E., et al., Detecting dementia with the mini-mental state examination in highly educated individuals. Archives of Neurology, 2008. 65(7): p. 963–967.

47. Gottesman, R.F., et al., Associations Between Midlife Vascular Risk Factors and 25-Year Incident Dementia in the Atherosclerosis Risk in Communities (ARIC) Cohort. JAMA Neurology, 2017. 74(10): p. 1246–1254.

48. Cole, J.H., Multimodality neuroimaging brain-age in UK biobank: relationship to biomedical, lifestyle, and cognitive factors. Neurobiology of Aging, 2020. 92: p. 34–42.

49. Popescu, S.G., et al., A U-Net model of local brain-age. bioRxiv, 2021: p. 2021.01.26.428243.

