## Supplementary Materials for "Brain-age predicts subsequent dementia in memory clinic patients"

### 1 MRI acquisition parameters

All 1140 MRI scans were T1-weighted and obtained from the same 1.5T MRI scanner (scanner A) at the Department of Neuroimaging, Institute of Psychiatry, Psychology and Neuroscience, King’s College London, UK. The MRI acquisition parameters varied slightly with a total of 22 types which are presented in **Table S1** including the count for each type.

**Table S1: MRI acquisition parameter types**

| MRI 'type' | Count | voxel dimensions (mm) x*y*z | | | Repetition time | Echo Time | Matrix size x*y*z | | | Slice thickness | Slice gap | Flip Angle | FoV | Inversion Time |
| --- | --- | --- | --- | --- | --- | --- | --- | --- | --- | --- | --- | --- | --- | --- |
| 1 | 433 | 1.25 | 1.25 | 1.2 | 8.592 | 3.8 | 192 | 192 | 180 | 1.2 | 1.2 | 8 | 240 | 1000 |
| 2 | 221 | 1.25 | 1.25 | 1.2 | 8.976 | 3.8 | 192 | 192 | 166 | 1.2 | 1.2 | 8 | 240 | 600 |
| 3 | 173 | 1.25 | 1.25 | 1.2 | 9.184 | 3.9 | 192 | 192 | 166 | 1.2 | 1.2 | 8 | 240 | 600 |
| 4 | 148 | 1.25 | 1.5 | 1.2 | 8.536 | 3.772 | 192 | 160 | 180 | 1.2 | 1.2 | 8 | 240 | 1000 |
| 5 | 53 | 1.25 | 1.5 | 1.2 | 8.932 | 3.776 | 192 | 160 | 166 | 1.2 | 1.2 | 8 | 240 | 600 |
| 6 | 45 | 1.25 | 1.25 | 1.2 | 8.812 | 3.9 | 192 | 192 | 180 | 1.2 | 1.2 | 8 | 240 | 1000 |
| 7 | 37 | 1.25 | 1.5 | 1.2 | 9.14 | 3.876 | 192 | 160 | 166 | 1.2 | 1.2 | 8 | 240 | 600 |
| 8 | 15 | 1.25 | 1.5 | 1.2 | 8.752 | 3.872 | 192 | 160 | 180 | 1.2 | 1.2 | 8 | 240 | 1000 |
| 9 | 2 | 1.25 | 1.25 | 1.2 | 8.976 | 3.8 | 192 | 192 | 150 | 1.2 | 1.2 | 8 | 240 | 600 |
| 10 | 1 | 0.859375 | 1.14583333 | 1.5 | 12.308 | 5.312 | 256 | 192 | 124 | 1.5 | 1.5 | 20 | 220 | 450 |
| 11 | 1 | 0.859375 | 1.14583333 | 1.5 | 12.428 | 5.348 | 256 | 192 | 124 | 1.5 | 1.5 | 20 | 220 | 450 |
| 12 | 1 | 1.25 | 1.25 | 1.2 | 9.192 | 3.904 | 192 | 192 | 136 | 1.2 | 1.2 | 8 | 240 | 600 |
| 13 | 1 | 1.25 | 1.25 | 1.2 | 8.984 | 3.804 | 192 | 192 | 138 | 1.2 | 1.2 | 8 | 240 | 600 |
| 14 | 1 | 1.25 | 1.5 | 1.2 | 9.148 | 3.88 | 192 | 160 | 140 | 1.2 | 1.2 | 8 | 240 | 600 |
| 15 | 1 | 1.25 | 1.5 | 1.2 | 9.14 | 3.876 | 192 | 160 | 144 | 1.2 | 1.2 | 8 | 240 | 600 |
| 16 | 1 | 1.25 | 1.25 | 1.2 | 8.976 | 3.8 | 192 | 192 | 146 | 1.2 | 1.2 | 8 | 240 | 600 |
| 17 | 1 | 1.25 | 1.5 | 1.2 | 9.14 | 3.876 | 192 | 160 | 148 | 1.2 | 1.2 | 8 | 240 | 600 |
| 18 | 1 | 1.25 | 1.5 | 1.2 | 9.14 | 3.876 | 192 | 160 | 154 | 1.2 | 1.2 | 8 | 240 | 600 |
| 19 | 1 | 1.25 | 1.25 | 1.2 | 9.184 | 3.9 | 192 | 192 | 160 | 1.2 | 1.2 | 8 | 240 | 600 |
| 20 | 1 | 1.25 | 1.25 | 1.2 | 9.06 | 3.792 | 192 | 192 | 166 | 1.2 | 1.2 | 8 | 240 | 600 |
| 21 | 1 | 1.25 | 1.25 | 1.2 | 9.952 | 4.012 | 192 | 192 | 166 | 1.2 | 1.2 | 8 | 240 | 600 |
| 22 | 1 | 1.25 | 1.5 | 1.2 | 8.62 | 3.792 | 192 | 160 | 180 | 1.2 | 1.2 | 8 | 240 | 1000 |

### 2 Supplementary Results

*Note*: to minimize the problem of multicollinearity between age and age^2^ we orthogonalized these variables by using their polynomial terms. However, a caveat with polynomial terms is that these can lead to uninterpretable coefficients. Analyses versions excluding polynomial terms can be accessed via the analyses code (see Methods section in the main document for the github link).

**Table S2.1: Logistic Regression results: full sample**

|  | **n/N** | **OR (CI)** | ***p*-value** |
| --- | --- | --- | --- |
| age | 476/1140 | 313951558535.82 (556454000.99-279323678981220.72) | <0.0001 |
| age2 | 476/1140 | 0.06 (0.00-27.63) | 0.4017 |
| sex | 476/1140 | 0.66 (0.51-0.87) | 0.0033 |
| brainPAD | 476/1140 | 1.03 (1.02-1.05) | <0.0001 |
| MMSE | 476/1140 | 0.96 (0.94-0.98) | <0.0001 |
| normVol | 476/1140 | 0.00 (0.00-0.00) | <0.0001 |

**Table S2.2 Logistic Regression results: sensitivity analysis 1**

|  | **n/N** | **OR (CI)** | ***p*-value** |
| --- | --- | --- | --- |
| age | 66/249 | 227063486.08 (91257.10-9320118318770.18) | <0.0001 |
| age2 | 66/249 | 0.16 (0.00-91.01) | 0.6371 |
| sex | 66/249 | 0.42 (0.21-0.80) | 0.0095 |
| brainPAD | 66/249 | 1.06 (1.03-1.10) | 0.0011 |
| MMSE | 66/249 | 1.00 (0.95-1.05) | 0.8889 |
| normVol | 66/249 | 0.78 (0.00-406.24) | 0.9360 |

**Table S2.3 Logistic Regression results: sensitivity analysis 2**

|  | **n/N** | **OR (CI)** | ***p*-value** |
| --- | --- | --- | --- |
| age | 146/471 | 66297618.38 (45434.53-544872748403.84) | <0.0001 |
| age2 | 146/471 | 0.15 (0.00-106.97) | 0.6255 |
| sex | 146/471 | 0.69 (0.44-1.06) | 0.0940 |
| brainPAD | 146/471 | 1.02 (1.00-1.05) | 0.0544 |
| MMSE | 146/471 | 0.95 (0.77-1.16) | 0.5837 |
| normVol | 146/471 | 0.00 (0.00-0.04) | 0.0005 |

**Table S2.4 Survival analysis: Cox proportional hazards regression results: full sample**

|  | **n/N** | **HR (CI)** | ***p*-value** |
| --- | --- | --- | --- |
| age | 476/1140 | 1141868143.44 (6876147.18-189621138509.18) | <0.0001 |
| age2 | 476/1140 | 0.02 (0.00-1.60) | 0.0808 |
| sex | 476/1140 | 0.74 (0.61-0.90) | 0.0020 |
| brainPAD | 476/1140 | 1.03 (1.02-1.04) | <0.0001 |
| MMSE | 476/1140 | 0.98 (0.97-0.99) | <0.0001 |
| normVol | 476/1140 | 0.00 (0.00-0.02) | <0.0001 |

**Table S2.5 Survival analysis: Cox proportional hazards regression results: sensitivity analysis 1**

|  | **n/N** | **HR (CI)** | ***p*-value** |
| --- | --- | --- | --- |
| age | 66/249 | 4706979.46 (2238.05-9899541040.96) | <0.0001 |
| age2 | 66/249 | 0.02 (0.00-6.80) | 0.1951 |
| sex | 66/249 | 0.55 (0.32-0.95) | 0.0327 |
| brainPAD | 66/249 | 1.06 (1.02-1.09) | 0.0006 |
| MMSE | 66/249 | 1.00 (0.97-1.04) | 0.9015 |
| normVol | 66/249 | 0.84 (0.01-127.17) | 0.9468 |

**Table S2.6 Survival analysis: Cox proportional hazards regression results: sensitivity analysis 2**

|  | **n/N** | **HR (CI)** | ***p*-value** |
| --- | --- | --- | --- |
| age | 146/471 | 12188042.26 (24700.01-6014100603.80) | <0.0001 |
| age2 | 146/471 | 0.48 (0.00-61.00) | 0.7649 |
| sex | 146/471 | 0.83 (0.59-1.16) | 0.2674 |
| brainPAD | 146/471 | 1.03 (1.01-1.05) | 0.0006 |
| MMSE | 146/471 | 0.86 (0.74-1.02) | 0.0782 |
| normVol | 146/471 | 0.01 (0.00-0.11) | 0.0008 |
